# Accurate overall, uneven by patient: a benchmark and demographic audit of deep learning for 12 lead ECG classification on PTB-XL

**DOI:** 10.64898/2026.07.09.26357670

**Authors:** Ahmad Durre Rehman, Samavia Nazir

**Affiliations:** CMH Institute of Medical Sciences (CIMS), Multan, Pakistan; NeuralCodes; CMH Institute of Medical Sciences (CIMS), Multan, Pakistan

**Keywords:** electrocardiography, deep learning, PTB-XL, algorithmic fairness, subgroup analysis, benchmarking

## Abstract

Deep learning reads 12 lead electrocardiograms at close to expert level on public benchmarks, yet most reports give one accuracy figure for the whole test set and stop there. We trained three architectures that are standard in this field, a 1D ResNet, a convolutional network with a bidirectional LSTM, and a convolutional network with a bidirectional LSTM followed by a transformer encoder, on the PTB-XL dataset to classify the five diagnostic superclasses, and then looked at how each one performed across sex and age. On the held out fold all three reached a macro AUC near 0.92, in line with the strongest published results on this benchmark, and the simplest model, the 1D ResNet, was marginally the best at 0.9241. The averages hid a steady pattern. Every model scored lower for female patients than for male patients, and every model scored lowest for patients aged 80 and over, where the 1D ResNet fell to 0.8878 and the transformer to 0.8693. Adding complexity did not close either gap and slightly widened the gap by age. Overall accuracy on PTB-XL is close to solved for these model families, but the benefit is not shared evenly, and a single headline number hides the patients a model serves worst. We release the full stratified evaluation to support fairness aware reporting.

## 1. Introduction

The electrocardiogram is the most common cardiac test in the world, and deep learning now reads it well. On PTB-XL, the largest openly available clinical ECG dataset, convolutional and recurrent networks classify diagnostic categories at a level that approaches expert agreement, with the best reported models reaching a macro AUC around 0.925. Most studies, though, report one number for the whole test set and treat that as the result.

A single number can mislead in medicine. A model that averages well can still be weak for a particular group of patients, and if that group is older, or sicker, or simply less represented in the training data, the average will hide it. Medical imaging has already shown this plainly, with classifiers that look accurate overall yet underdiagnose women and patients from under served backgrounds. ECGs have had far less of this scrutiny, although the same risk applies.

We take three architectures that are standard in this area and put them through one identical pipeline on PTB-XL, then look past the average. The aim is not a new network. It is to ask a plain question that the usual reporting skips: once these models are accurate on the whole test set, are they equally accurate for men and women, and for the young and the old.

We report four things. We give a like for like comparison of a 1D ResNet, a CNN with a bidirectional LSTM, and a CNN with a bidirectional LSTM followed by a transformer, trained and evaluated the same way. We show that all three reach a macro AUC near 0.92, matching the published benchmark, and that the simplest of them is marginally the best. We then show a consistent demographic pattern beneath those averages, with lower performance for female patients and, more sharply, for the oldest patients. Finally we show that adding complexity did not close these gaps and widened the age gap, which is worth stating on its own.

## 2. Background

PTB-XL set a reference point for this task. The dataset provides more than twenty thousand clinical 12 lead recordings with expert labels, and the accompanying benchmark showed that ordinary convolutional networks classify the diagnostic superclasses at a macro AUC near 0.925, with deeper or fancier designs offering little beyond that. A recurring lesson from that work is that simple convolutional models are already very strong here, so accuracy alone no longer separates architectures by much.

Fairness in clinical machine learning is a separate and growing concern. Studies of chest radiograph classifiers found that models with high overall accuracy still missed disease more often in female patients and in groups that were under represented in training, and the response has been a push toward reporting performance for each subgroup rather than for the pooled test set. ECG models have not been examined this way as often, even though age and sex both shape the ECG signal and both are recorded in PTB-XL. That is the gap this study addresses.

## 3. Methods

### Dataset

We used PTB-XL, which contains 21799 records from 18869 patients, each a ten second 12 lead recording. We used the 100 Hz version of the waveforms. Labels were reduced to the five diagnostic superclasses, normal (NORM), myocardial infarction (MI), ST and T wave changes (STTC), conduction disturbance (CD), and hypertrophy (HYP), using the standard mapping supplied with the dataset, and treated as a multilabel problem because a record may carry more than one. We followed the official ten fold split, training on folds one to eight, validating on fold nine, and testing once on fold ten.

### Preprocessing

Each lead was standardized using the mean and standard deviation of the training set, and the same statistics were applied to the validation and test sets. No other filtering was applied.

### Models

The three networks are established building blocks rather than novel designs, which is deliberate, since the point is the comparison and the audit, not a new architecture. The 1D ResNet is a stack of residual convolutional blocks with a global average pool and a linear head. The CNN BiLSTM passes a short convolutional stack into a bidirectional LSTM, averages over time, and predicts with a linear layer. The CNN BiLSTM Transformer adds a transformer encoder after the recurrent layer before pooling. All three take a twelve by one thousand input and output five scores.

### Training

Every model was trained with the same recipe, binary cross entropy on the five outputs, the Adam optimizer at a learning rate of 0.001, a batch size of 128, for twenty epochs, with the checkpoint that scored the best validation macro AUC kept for testing. A fixed random seed was used throughout.

### Evaluation

The primary measure was the macro average of the area under the receiver operating characteristic curve across the five classes on the test fold. We also report the area under the curve for each class, and, for the fairness audit, the macro AUC computed separately within each subgroup. Subgroups were sex, as recorded in the dataset, and four age bands, under 50, 50 to 65, 65 to 80, and 80 and over. A subgroup with too few cases to score a class was excluded from that class. Reporting followed the TRIPOD+AI guidance. Because the dataset is public and the training recipe is fixed, the study is straightforward to reproduce.

## 4. Results

### Overall accuracy

All three models classified the five superclasses at a macro AUC near 0.92 on the held out fold, and the three sat within 0.005 of each other, so on accuracy alone they are hard to separate. The simplest model, the 1D ResNet, was marginally the best at 0.9241. These figures match the published PTB-XL benchmark closely, which is the clearest sign that the pipeline is sound and the numbers are trustworthy. Across classes, normal recordings were the easiest and hypertrophy the hardest, a pattern shared by all three models. Table 1 gives the full breakdown.

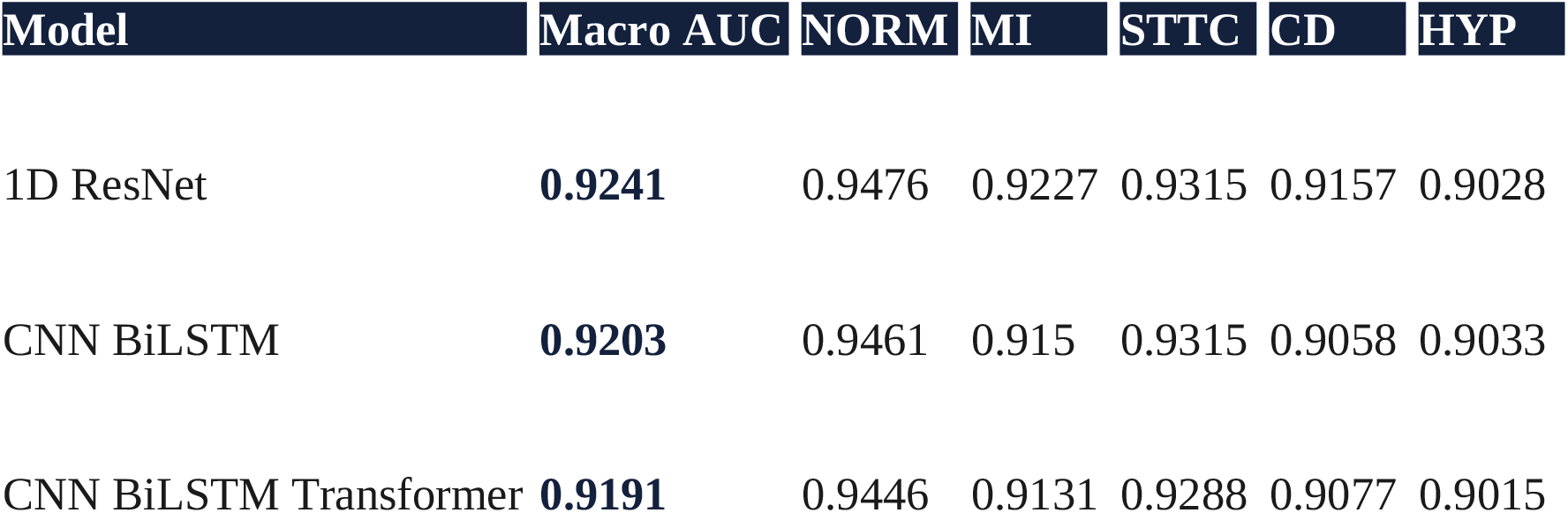

### Performance by subgroup

The averages concealed a consistent split, shown in Table 2 and Figure 2. On sex, every model scored higher for male patients than for female patients, by between 0.0148 and 0.0177 points of macro AUC, a small gap but one that pointed the same way in all three models and grew slightly as the model became more complex. On age the effect was larger and just as consistent. Every model scored lowest for patients aged 80 and over. For the 1D ResNet the macro AUC fell from 0.9219 in the under 50 group to 0.8878 in the oldest group, a drop of 0.0341. For the transformer the same comparison fell from 0.9217 to 0.8693, a drop of 0.0524, the largest of the three.

**Figure 1.**
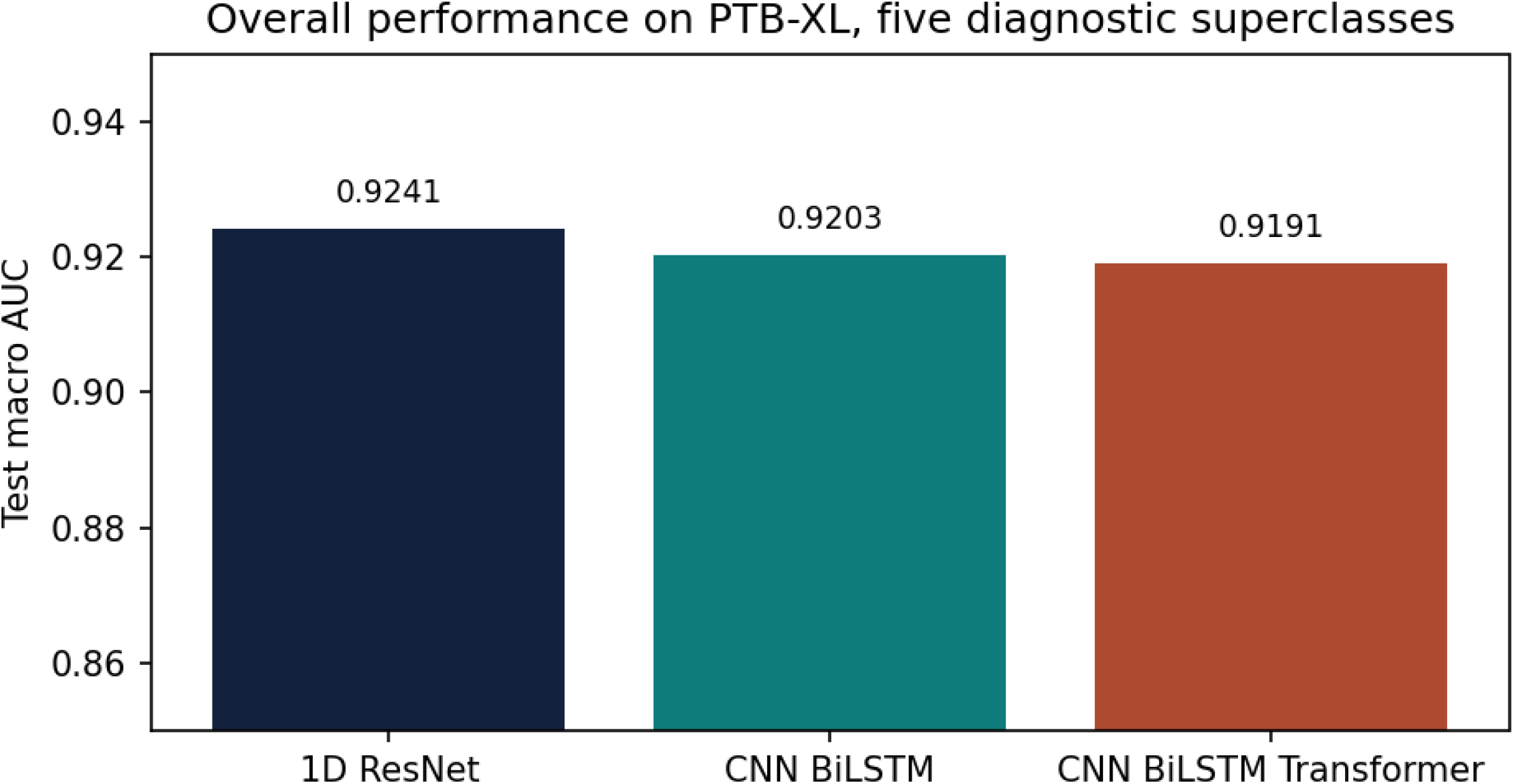
Overall macro AUC on the PTB-XL test fold for the three architectures.

**Figure 2.**
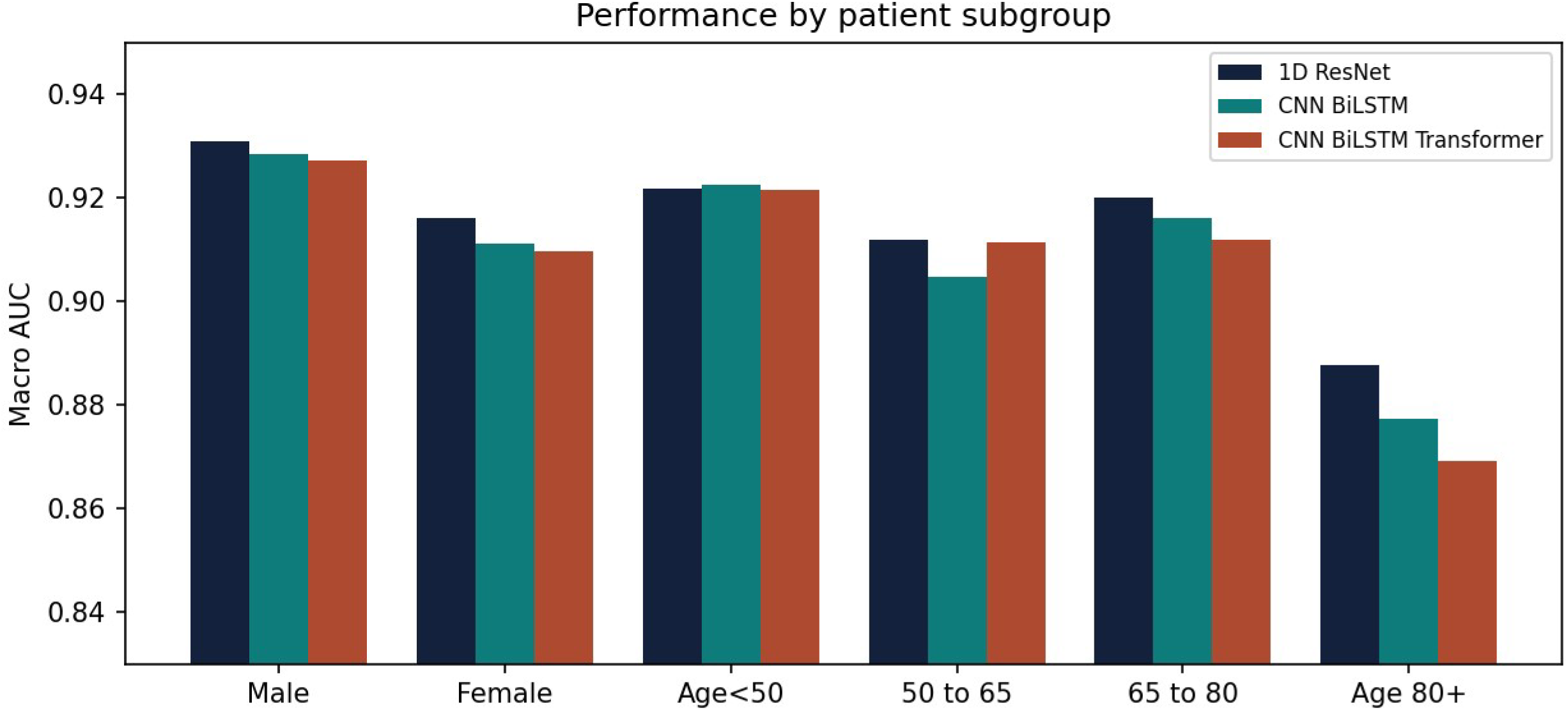
Macro AUC within each patient subgroup. Every model is lowest for the oldest patients, and the transformer shows the widest age gap.

| Model | Male | Female | Sex gap | Age <50 | 50 to 65 | 65 to 80 | Age 80+ | Age gap |
| --- | --- | --- | --- | --- | --- | --- | --- | --- |
| 1D ResNet | 0.9309 | 0.9162 | 0.0148 | 0.9219 | 0.912 | 0.9201 | 0.8878 | 0.0341 |
| CNN BiLSTM | 0.9286 | 0.9113 | 0.0173 | 0.9226 | 0.9048 | 0.9162 | 0.8773 | 0.0453 |
| CNN BiLSTM Transformer | 0.9274 | 0.9097 | 0.0177 | 0.9217 | 0.9114 | 0.9119 | 0.8693 | 0.0524 |

The relationship between complexity and fairness ran the wrong way. The extra capacity of the recurrent and transformer models bought no gain in overall accuracy over the plain 1D ResNet, and the transformer showed the widest gap for the oldest patients. Bigger did not mean fairer here.

## 5. Discussion

Two findings sit side by side. Accuracy on PTB-XL is close to saturated for these model families, and the simplest network matches the more elaborate ones, which agrees with earlier work and means architecture choice is not where the remaining problem lies. The remaining problem is distributional. The oldest patients, and to a smaller degree female patients, were served worse by every model we tried. Older ECGs tend to be noisier and to carry more mixed pathology, which is a plausible driver of the drop, but a model deployed without awareness of this would fail precisely the group at highest cardiac risk. The finding that the transformer widened rather than narrowed the age gap is a caution against the assumption that larger models are automatically fairer. Here they were not.

The practical message is simple. Report performance for each subgroup, not only a pooled figure, because the pooled figure can look excellent while a model quietly underperforms for a group that matters. The audit also argues for validating a model on the population it will actually serve. PTB-XL is drawn largely from one European cohort, and a South Asian or Pakistani population is barely represented in public ECG data. We are pursuing that external validation on a local cohort as the next step, and this benchmark is the baseline it will be measured against.

This study has limits that bound its claims. It uses one dataset, so the numbers describe PTB-XL and not ECG classification in general. It compares three architectures rather than an exhaustive set, although those three span the common families. Sex is recorded as a binary in the source data, the age bands are coarse, and there is no external test set here. Each of these is a direction for the work that follows.

## 6. Conclusion

Three standard deep learning models read the PTB-XL diagnostic superclasses at a macro AUC near 0.92, but not equally for everyone. The oldest patients were consistently underserved, female patients slightly so, and adding model complexity did not fix either gap. Aggregate accuracy is not enough. Stratified reporting should be the default for ECG models, and validation on the intended population should follow.

## Data Availability

https://physionet.org/content/ptb-xl/1.0.3/
https://github.com/ahmaddurrerehman-glitch/ecg-ptbxl-fairness

https://github.com/ahmaddurrerehman-glitch/ecg-ptbxl-fairness

## Data and code availability

PTB-XL is publicly available under a Creative Commons Attribution license from PhysioNet. The training and evaluation code is available at https://github.com/ahmaddurrerehman-glitch/ecg-ptbxl-fairness.

## Ethics

This work is a secondary analysis of the publicly available, deidentified PTB-XL dataset. It involves no new human subjects data and required no additional ethical approval. The original dataset was released with institutional ethics approval.

## Author contributions

A.D.R. designed the study, implemented and trained the models, ran the analysis, and drafted the manuscript. S.N. provided the clinical interpretation of the diagnostic categories and the subgroup findings, advised on the clinical framing and significance of the fairness analysis, and reviewed and revised the manuscript for clinical accuracy. Both authors reviewed and approved the final manuscript.

